# An efficient design for whole genome trio sequencing identifies key variants in rare neurological disorder cases

**DOI:** 10.1101/2023.10.13.23296768

**Authors:** K. Ramsey, S. Kruglyak, M. Naymik, B. R. Lajoie, K. N. Wiseman, M. Sanchez-Castillo, S. Billings, W. Jepsen, M. Huentelman, V. Narayanan

## Abstract

We sequenced nine trios in which the probands in an underserved population were affected by a rare and undiagnosed disorder with neurological features. Sequencing was performed with one trio per flowcell on a benchtop sequencing instrument, leveraging the design of sequencing the proband at twice the coverage of the parents. The reduced coverage in the parents led to sequencing efficiencies while retaining the benefits of trio sequencing: the ability to discover de novo variants and the ability to trace inheritance patterns of rare variants. Once the sequencing data was generated, our two teams used independent informatics pipelines for variant calling and interpretation. In five of the nine cases, both teams found a single SNV or small indel that was deemed causal pending clinical validation. In three of the nine cases, neither team had a significant finding. In the final case, an additional scan for large CNVs performed by one of the teams identified a de novo deletion and duplication in the proband which is the likely cause of the underlying disease. The results across cases with significant findings showed a variety of affected genes (CFAP52, DYNC1H1, FANCE, TCF4, and TOP3A), variant types, and inheritance patterns. All findings were clinically validated, after which the families were counseled about disease management, current research studies (i.e. gene therapy), and family planning. With six of nine families receiving findings, the study demonstrated an efficient and effective trio sequencing design strategy.

## Introduction

Whole genome sequencing is a powerful tool for discovering causal variants in rare disease studies^1–3^. In previous work, whole genome sequencing by avidity effectively identified causal variants in approximately half of the genomes sequenced in cases of rare eye disease^4^. The work primarily focused on single genomes and all findings involved inherited SNPs or small indels. In this study, we sequenced nine trios consisting of a proband affected by a rare, undiagnosed neurological disorder and both parents to enable identification of *de novo* variants, which are often the cause of genetic neurological disorders^5^. We chose to sequence the proband at a typical coverage level of approximately 35-40X and the parents at half that coverage (∼15-20X). The design is efficient in cost and time because the three genomes can be sequenced in a single flowcell of the AVITI^6^ benchtop sequencing instrument. We hypothesized that we would be able to observe inheritance patterns and identify *de novo* variants even with the reduced coverage in the parents.

The families for the study were selected from an underserved population and enrolled at TGen’s Center for Rare Childhood Disorders. The families had little to no access to previous genetic testing. Enrollment in this study provided a means of finding the underlying cause of their child’s disorder and hope in ending the diagnostic odyssey. The probands’ symptoms included global developmental delay, intellectual disability (ID), seizures, hypotonia, microcephaly, situs inversus, facial dysmorphism, thrombocytopenia, small stature, and cardiac malformations (see Table 1 for a complete list of phenotypes).

**Table 1:** Findings across all cases. Coordinates are provided with respect to the hg19 / GRCh37 reference.

| ID | Gene | Variant | Inheritance | Phenotypes | TGen Recommendations |
| --- | --- | --- | --- | --- | --- |
| 1 | DYNC1H1 | c.4580delinsCC, p.L1527Pfs*9 | Het *de novo* | Global developmental delay, ID, Seizures, Cerebral hemorrhage | Referred family to support group, research, discussed recurrence risk |
| 2 | TCF4 | c.923-2 A>G, splice acceptor variant | Het *de novo* | Global developmental delay, Nonverbal, Hypotonia, Short stature, Happy demeanor | Referred family to the support group and registry at the Pitt Hopkins Research Foundation, gene therapy and therapeutic drug research, discussed recurrence risk |
| a3 | ChrX terminal del/dup | Terminal deletion 1.45 Mb Xp22.33 and terminal duplication 15.1 Mb Xq27.1 to Xq28 | Het *de novo* | Developmental delay, Microcephaly, Hypotonia, Hypoplasia of corpus callosum, High pain tolerance, Seizures, Hypothyroidism, Facial dysmorphism, Cryptorchidism and Micropenis | Provided resources from Unique, recommendations of FISH studies in at least mother but also proband's siblings to determine if mother has a pericentric inversion |
| 4 | CFAP52 | c.1162del p.S388Afs*21 | Homo recessive | Global developmental delay, Hearing impairment, Dextrocardia, Transposition of the great arteries, Situs inversus totalis | Recommendations for creating social media support group, family planning |
| 5 | TOP3A | c.1685 G>A, p.G562E | Homo recessive | Global developmental delay, Microcephaly, Short stature, Decreased body weight, Periventricular leukomalacia, Delayed gross motor development | Recommended sister chromatid exchange studies, follow-up ECHO, discussion of "mitochondrial cocktail" with her healthcare team, discussed recurrence risk |
| 6 | FANCE | c.1111 C>T, p.R371W | Homo recessive | Thrombocytopenia, VSD, Single kidney, Unilateral deafness, R Thumb abnormality, Short stature, Triangular face, Narrow nasal bridge, Learning delays | Referral to the Fanconi Anemia Research Fund and Family Support Group, discussion of hematopoietic cell transplant, cancer surveillance, family planning |
| 4 | PALB2 | c.2257 C>T, p.R753* | Het maternal | Incidental Finding: Risk for Hereditary Breast and Ovarian Cancer (HBOC) | Counseled mother about risk for HBOC, recommendations for increased screening, risk reducing surgery, testing of family members; mother does not meet NCCN criteria for testing based on personal and family history |

## Results

Extracted DNA was used to make PCR-free sequencing libraries for the 27 samples (9 affected probands and their parents). The libraries were pooled by trio, with each proband having twice the concentration as the parents. Following whole genome sequencing and FASTQ generation, the data was independently analyzed by the teams at Element Biosciences and at TGen. We wanted to determine whether teams working independently and with different informatics pipelines would arrive at the same interpretations for each of the cases. One team used a combination of DNAScope (Sentieon, San Jose California) and Franklin (Genoox, Tel Aviv, Israel) for secondary and tertiary analysis. The other team leveraged Varseq (Golden Helix, Inc., Bozeman, MT) and Emedgene (Illumina, San Diego, CA) tools. Once interpretation was completed, final results were shared and discussed. In five of the cases, the two teams identified the same variant as causal. In 3 of the cases, neither team identified a causal variant. In the final case, one team identified a large CNV as the likely cause of the disorder, while the other team did not have CNV interpretation implemented within their pipeline. Following data review, both teams agreed on the six findings and samples were sent for clinical validation either via Sanger sequencing in a CLIA lab (for SNPs and small indels) or for CMA in a CLIA lab for the large CNV (GeneDx, Gaithersburg, MD). All variants were confirmed, at which point results could be shared with the treating clinicians and family counseling could begin. Table 1 summarizes the findings.

In three cases (Families 1, 2, and 3), *de novo* variants were determined to be causal. One of the variants was an indel in the *DYNC1H1* gene and the other was a splice acceptor variant in *TCF4*. Figure 1 shows the IGV displays for the two calls and demonstrates how the trio design was leveraged to confirm that neither variant is inherited.

**Figure 1:**
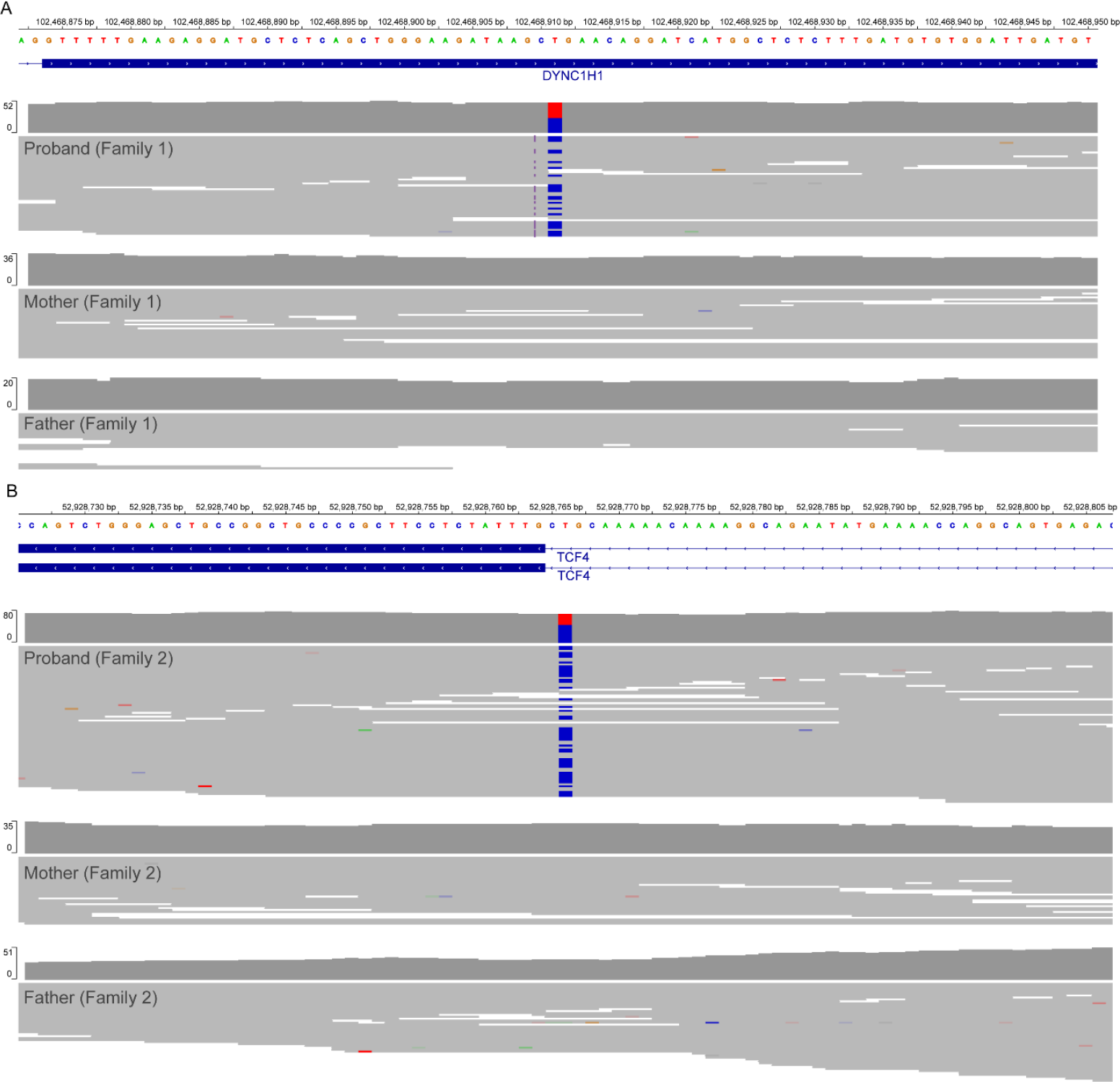
(A) De novo indel in DYNC1H1 gene. The proband is in the top display and the parents are below. The reference T is replaced by a CC, causing a heterozygous frameshift. The data from the parents shows no evidence of the variant, confirming that the mutation occurred de novo. (B) Splice altering de novo variant in TCF4. The top display in the proband confirms the heterozygous variant, while the maternal and paternal displays below show no evidence for the variant in the parents, confirming the de novo call.

In one of the cases (Family 3), a del/dup event in chromosome X (chrX) was discovered. WGS did not detect a chromosomal rearrangement or del/dup on chrX in the mother. Figure 2 shows the structural variants in chrX in the trio.

**Figure 2:**
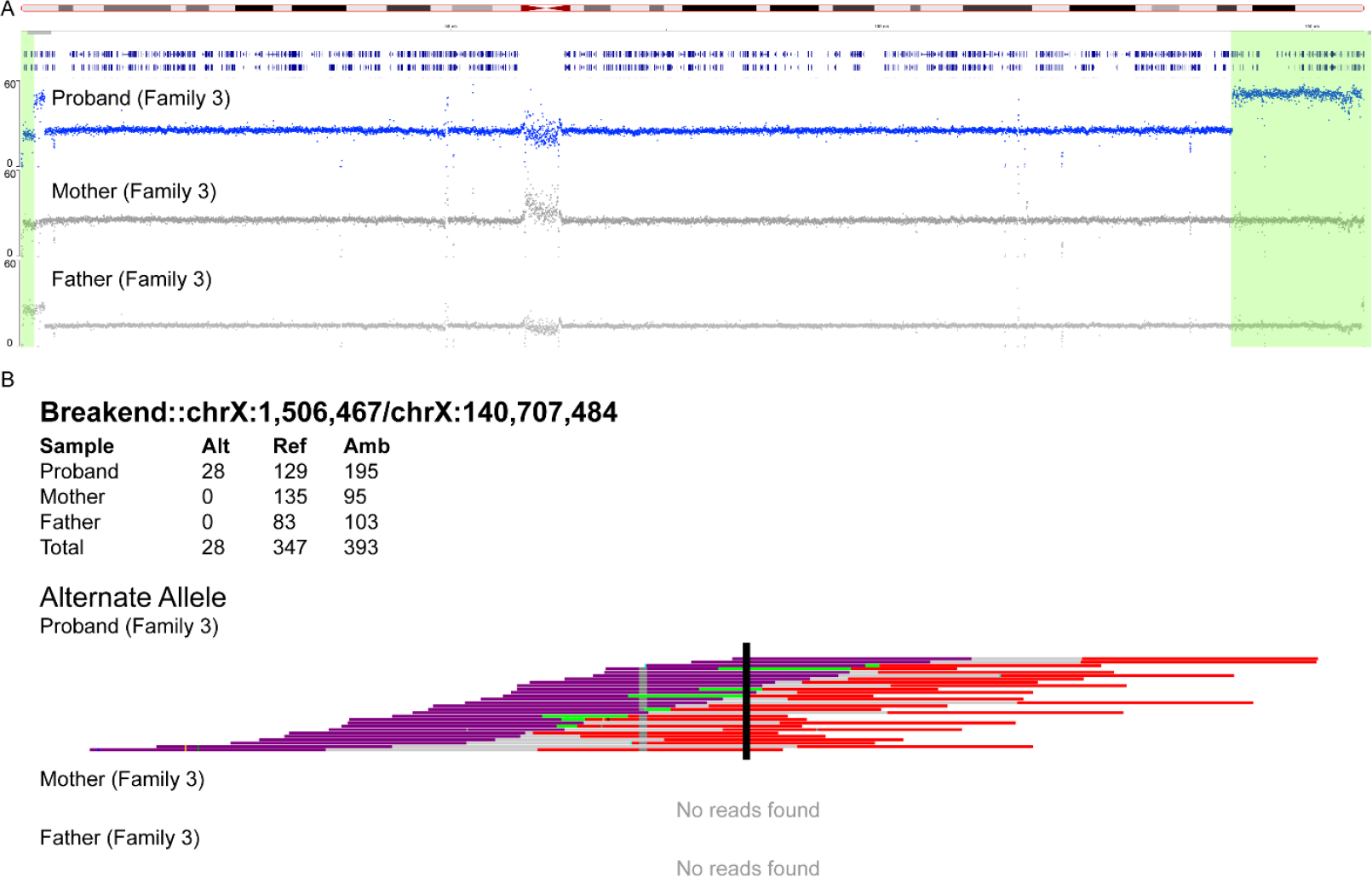
(A) Chromosome X coverage in the proband (top) followed by the mother and father. The proband has a deletion in the beginning of the chromosome marked in green. This is part of the PAR region so a copy number of 2 is expected in males, as shown in the father, but the proband has a stretch of copy number 1. The proband also has a large duplication at the end of the chromosome. (B) SVVIZ7 display of the junction between the duplication and an early part of the chromosome. The proband shows extensive read support for the junction, while the parents show no support. This leads to the conclusion that the event is de novo.

In three cases (Families 4, 5, and 6), the primary finding was a small homozygous variant in the proband (in genes *CFAP52, TOP3A*, and *FANCE*, respectively). Surprisingly, each allele was extremely rare in the population, yet the proband inherited a copy from each of the unrelated parents. Intuition would suggest that a different rare allele would be inherited from each parent. Table 2 provides the gnomAD allele frequencies and the relatedness coefficients^8^ for each case. The rare nature of these variants and the cooccurrence in a single family may imply the presence of a founder mutation or a ‘mutation hotspot.’

**Table 2:** Allele frequencies and relatedness coefficients for the three homozygous variants. Surprisingly, each parent is a carrier for the same rare variant. Coordinates are provided with respect to the hg19 / GRCh37 reference.

| Gene | Variant | gnomAD het frequency | Parent relatedness coefficient |
| --- | --- | --- | --- |
| *CFAP52* | c.1162del<br>p.S388Afs*21 | 0.00006572 | 2.4% |
| *TOP3A* | c.1685 G>A, p.G562E | 0 | 2.3% |
| *FANCE* | c.1111 C>T, p.R371W | 0.00003288 | -2.0% |

## Discussion

In this study, we used a novel trio sequencing design to solve six of nine rare neurological disorder cases. Each trio was sequenced on a single flowcell of a benchtop sequencer, obviating the need for batching large numbers of samples. The design of sequencing parents at half the typical depth was effective in identifying *de novo* variants. The design also decreased manual interpretation effort (a costly step of the entire process) by dramatically shortening the list of variants prioritized by the automated machine learning algorithms. This is because rare heterozygous variants inherited from a healthy parent could be ruled out.

Both families 1 and 2 were counseled of a less than 1% recurrence risk (due to germline mosaicism). Both families were informed of online social media groups for the disorders, and Family 2 was directed to the Pit Hopkins Research Foundation (https://pithopkins.org/), which has support for families, a registry, and a list of currently funded research, including gene therapy and repurposing of drug compounds. For Families 4, 5, and 6, additional counseling was provided regarding recurrence risk and family planning for autosomal recessive disorders. Two of the families were interested in having additional children and were counseled on the recurrence risk of 25%, as well as the availability of PGD testing, sperm or egg donation, and adoption.

In Family 3, WGS identified a del/dup of chromosome X in the proband. Similar del/dups of chromosome X have been seen in individuals after a meiotic recombination event involving a phenotypically normal mother with a pericentric inversion on the X chromosome ^9,10^. However, a thorough analysis of both maternal and paternal WGS shows no indication of an inversion, suggesting the event is *de novo* in the proband. Confirmation studies using FISH would help support this finding, given other unexplained conditions in the pedigree (Figure 3).

**Figure 3:**
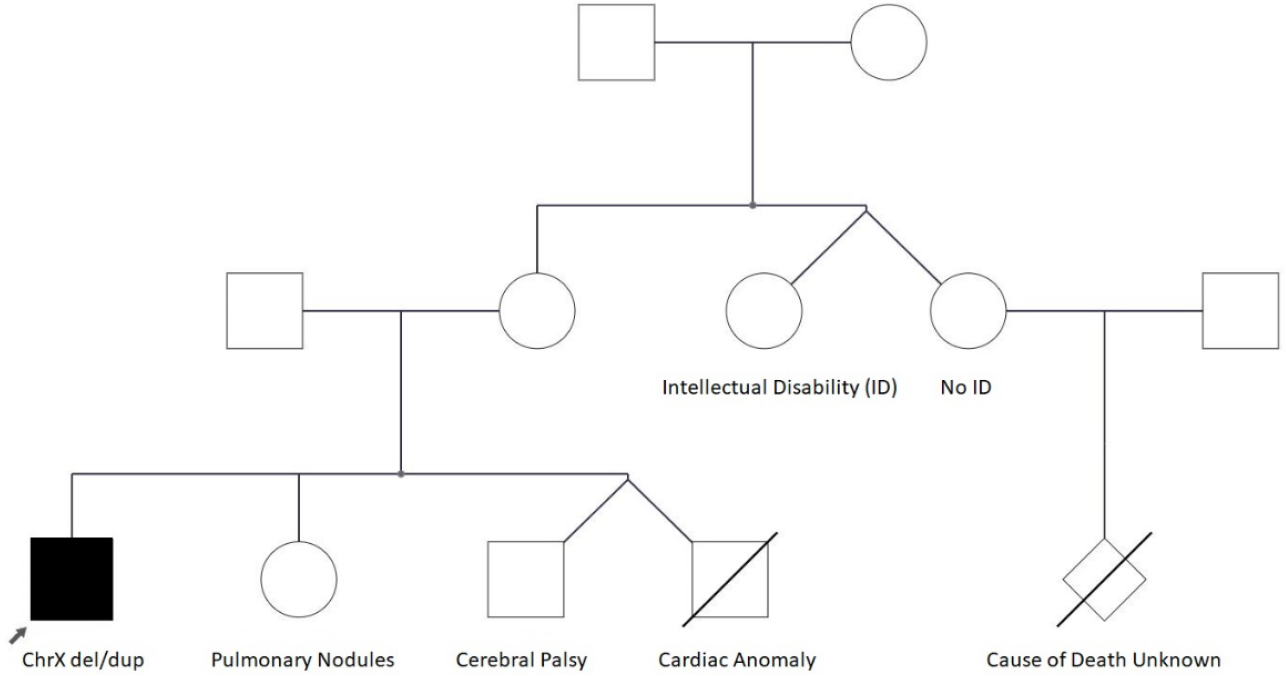
Pedigree of family 3. We had suspected a pathogenic variant segregating in the pedigree because multiple individuals are affected. However, we found the del / dup to be de novo and the conditions in the pedigree are highly varied.

However, at this time, it has not been possible to coordinate additional testing in the mother or siblings. The duplicated region contains more than 130 genes, including the *MECP2* gene. Duplications of 0.3 Mb and larger in *MECP2* are associated with *MECP2* duplication syndrome, causing early-onset hypotonia, ID, autistic features, seizures, spasticity, and lack of speech (Van Esch, 2008). The deleted region of the X chromosome contains ten genes, including *SHOX* and *CSF2RA*. The phenotype of *SHOX* deficiency disorders includes short stature, limb anomalies, including mesomelia and Madelung deformity^11^. Variants in the *CSF2RA* gene have been associated with pulmonary alveolar proteinosis ^12^. The proband’s phenotype overlaps with individuals with *MECP2* duplication syndrome as well as other individuals with similar chromosome X del/dup ^9,10,13^.

The proband in Family 5 was found to have a homozygous variant in *TOP3A* associated with a Bloom syndrome-like disorder ^14^. Unlike Bloom syndrome, individuals with *TOP3A* variants have shown evidence of mitochondrial dysfunction and dilated cardiomyopathy (sometimes fatal in childhood). Our patient had a normal echocardiogram at birth and is no longer being followed by cardiology. We recommended that since our patient was now older, she receive a follow-up echocardiogram. Based on the nature of the disorder, we also suggested the patient discuss a trial of “mitochondrial cocktail” (supplements including vitamins, Coenzyme Q10, and L-carnitine) with her healthcare providers.

The proband for Family 6 presented to our center with a recent diagnosis of thrombocytopenia and a history of short stature, cardiac anomaly, a single kidney, unilateral deafness, and a smaller and inflexible right thumb. The geneticist suspected Fanconi anemia (FA) or Russell Silver syndrome. Genetic testing was crucial in confirming the diagnosis as well as identifying the mode of inheritance—FA is associated with 21 genes and has autosomal recessive, dominant, and X-linked modes of inheritance ^15^. Beyond the traditional suggested cancer screening for FA, there is additional cancer surveillance recommended for individuals with mutations in genes associated with hereditary breast and ovarian cancer (HBOC), making a genetic diagnosis crucial for management. The proband was found to have a mutation in *FANCE*, a gene that accounts for only 3% of FA. The family was counseled about treatments to improve blood counts and the use of hematopoietic stem cell transplantation. The family was also referred to the Fanconi Anemia Research Fund for support, registry information, and current research and clinical trials (https://www.fanconi.org/).

In Family 4, a pathogenic variant in the PALB2 gene was found in both the proband and his mother. They had consented to receiving incidental findings. The family was provided counseling for an increased risk of HBOC and the mother was encouraged to notify her health care providers to establish increased screening and discuss surgical recommendations. The proband’s mother has no personal or family history of cancer and would not have met NCCN criteria for testing^16^. This case demonstrates the importance of universal screening for HBOC.

## Conclusion

We implemented an efficient trio sequencing design for the study of rare disorders and used it to find key variants in 6 of 9 cases. The findings included multiple variant types, including SNPs, indels, and structural variants. In three solved cases, the variants in the proband arose *de novo*, whereas in the other three cases, they were inherited. In addition to the primary findings, there was an incidental finding: a SNP conferring cancer risk. Although the two teams performing interpretation used different informatics tools for secondary and tertiary analysis, the findings matched closely. The only differences were encountered when one team used methods (e.g. CNV detection) that were not implemented by the other team. However, whenever each team had a finding for a case, the finding matched. All findings were clinically confirmed, and results were then returned. While having a diagnosis was already a positive result for the families in ending the diagnostic odyssey and suggesting support groups for the identified disorders, in some cases additional actions pertaining to family planning or clinical management were taken. For example, in the case of *de novo* variants, the families were told that recurrence risk in subsequent pregnancies would be minimal. By contrast, the risk in the cases where both parents were determined to be carriers for recessive disorders, the recurrence risk is 25%. Furthermore, certain findings suggested additional tests (e.g. ECHO cardiogram in the TOP3A case) or evaluation of potential gene therapy trials (e.g. the TCF4 case). We believe that the trio sequencing design described in this article could easily be scaled to more cases and effectively used in the study of rare genetic disorders.

## Methods

### Patient recruitment

Five male and four female probands, in the age range from 1-20 years, were either self-referrals to the Center for Rare Childhood Disorders or were referred by their neurologist or geneticist. Written informed consent for sequencing, data usage, and publication of clinical details was obtained from participants, legally authorized representatives and/or their guardians if under the age of 18 for all of the family members. The study protocol and consent procedure were approved by the WGC Institutional Review Board (WGC IRB; study number 20120789).

Whole blood was obtained for all participants except for one proband, where a buccal swab was obtained.

### Library Preparation

Libraries were prepared PCR-free using 500ng of input gDNA using the KAPA HyperPrep kit. The libraries were sheared on Covaris ME220 to ∼350bp average fragment size. KAPA UDI adapters were ligated and the libraries were size-selected with a dual-sided SPRI (0.48X/0.64X). All PCR-free libraries were circularized with the Element Adept Library Compatibility Kit v1.1 (cat# 830-00007). Following qPCR, the libraries were normalized to 1nM and pooled by volume as follows: proband 50%, mother 25%, and father 25%.

### Sequencing

Pooled libraries were shipped to FYR Diagnostics (Missoula, MT) for sequencing with 2×150 bp reads and a trio per flowcell on an Element Biosciences AVITI sequencer. Table 3 summarizes the coverage and quality score statistics for each trio. FASTQ files were shared with the Element Biosciences team and the TGen team.

**Table 3:** Summary of data collected and quality for each of the 27 samples in the study. Number following F is the deidentified family ID and the underscore 1, 2, 3, refers to proband, mother, and father, respectively.

| Sample Identifier | Data generated (GB) | %Q30 |
| --- | --- | --- |
| F1_1 | 130 | 95.3 |
| F1_2 | 62 | 95.7 |
| F1_3 | 53 | 95.6 |
| F2_1 | 166 | 94.2 |
| F2_2 | 70 | 95 |
| F2_3 | 78 | 94.7 |
| F3_1 | 164 | 94.7 |
| F3_2 | 78 | 94.7 |
| F3_3 | 87 | 95 |
| F4_1 | 154 | 95.3 |
| F4_2 | 87 | 95.2 |
| F4_3 | 79 | 94.9 |
| F5_1 | 160 | 95.4 |
| F5_2 | 74 | 95.1 |
| F5_3 | 83 | 95.9 |
| F6_1 | 159 | 94 |
| F6_2 | 84 | 95.5 |
| F6_3 | 83 | 95.2 |
| F7_1 | 113 | 92.3 |
| F7_2 | 61 | 93 |
| F7_3 | 75 | 94.3 |
| F8_1 | 164 | 94.6 |
| F8_2 | 79 | 94.6 |
| F8_3 | 87 | 94.6 |
| F9_1 | 171 | 94.6 |
| F9_2 | 82 | 94.1 |
| F9_3 | 78 | 94.3 |

### Element Biosciences Analysis

BWA-MEM was used to align the FASTQ files to the hg38 / GRCh38 human reference. Sentieon DNA scope was used for variant calling in each individual. VCF files and phenotypes were uploaded into the Franklin by Genoox interpretation software. Each prioritized variant was manually reviewed for quality, inheritance mode, and phenotypic fit. In five of the cases, a single variant was identified. For all cases, we also reviewed a coverage depth by chromosome plot. In one of the cases, we noticed that chromosome X had slightly higher coverage in the proband (Figure 4).

**Figure 4:**
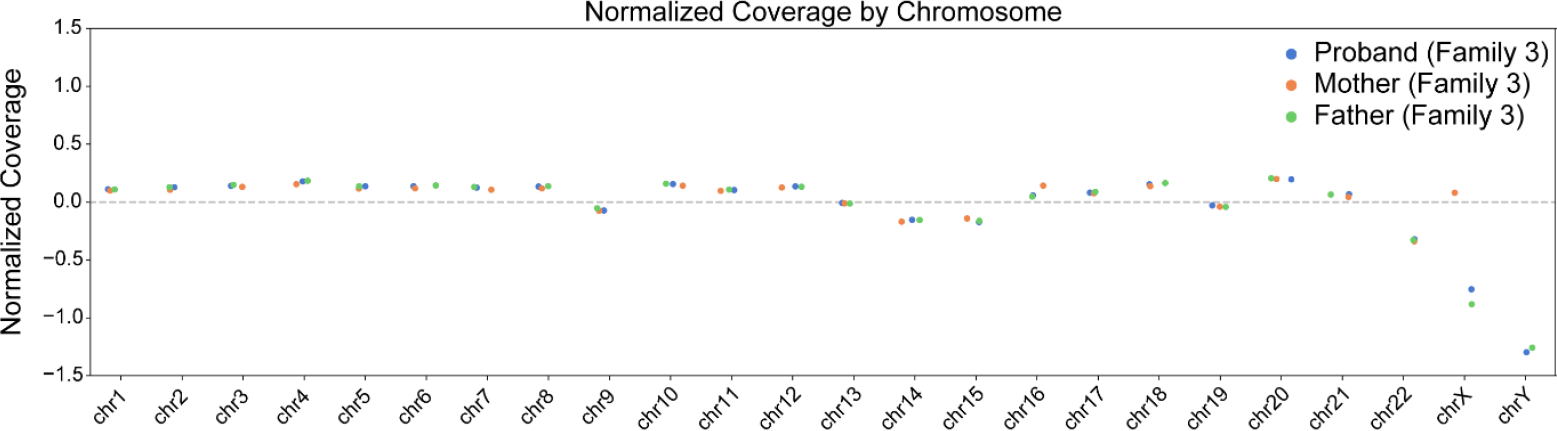
Normalized coverage by chromosome plot for the pedigree. The small increase in coverage in chromosome X of the proband relative to the father led to the hypothesis that a large CNV might be present.

A review in IGV revealed a large duplication. At this point, we uploaded the FASTQ files into the Franklin software and repeated analysis of all cases because Franklin analysis from FASTQ includes structural variant calling. The five prior findings were reproduced, but there were two additional findings. In the case where we had observed the duplication, several structural variants were found, including a deletion near the beginning of chromosome X and the duplication at the end of chromosome X. To determine that the del dup originated *de novo*, we entered the breakpoints into SVVIZ^7^ and observed clear evidence in the proband but no evidence in either parent. For three of the cases, there were no significant findings.

### TGen Analysis

Reads were aligned to the human genome (hg19/GRCh37) using the Burrows-Wheeler Aligner (BWA mem v.0.7.8). PCR duplicates were identified using Picard MarkDuplicates v1.79. Base quality recalibration and indel realignment were performed using the Genome Analysis Toolkit (GATK v3.5-1). Variants were jointly called with HaplotypeCaller and recalibrated with GATK. Quality controls were conducted using FASTQC v0.11.5. Called variants were annotated with SnpEff v3.0a against Ensembl GRCh37.66 and filtered against dbSNP137 1000 Genomes Project (minor allele frequency < 0.05), SnpEff Impact: High + Moderate, GATK quality score > 40, and known genes. Final annotation and reports were generated with Varseq (Golden Helix, Inc., Bozeman, MT) and Emedgene (Illumina, San Diego, CA). Relatedness coefficients used in Table 2 were part of the Emedgene analysis output.

### Review of interpretation results

Once both teams completed interpretation of the cases, we held a conference call to discuss the results. For five of the cases with findings, both teams identified the same variant. For three of the cases, neither team had a compelling finding. In the case of the del dup on chromosome X, only one team ran a structural variant caller and saw the event. However, during the meeting, the teams reviewed the findings in IGV, noted the genes in the affected region, and agreed that this was likely causal, pending clinical validation.

### Clinical validation

DNA samples were sent for clinical validation in a CLIA lab either via Sanger for SNPs and small indels or for chromosomal microarray in a CLIA lab for the large CNVs (GeneDx, Gaithersburg, MD).

## Data Availability

Data is available upon request subject to privacy restrictions

